# Insights from a double-blind, randomized, direct-to-participant intervention trial for Long COVID

**DOI:** 10.64898/2026.08.19.26360832

**Authors:** Julia Moore Vogel, Janna Ter Meer, Romina Foster-Bonds, Mario Patrice Duff, Andrea Goosen, Anastasia Kurakova, Ethan Dinh-Luong, Lena Miyasaki, Sarah Topol, Cynde Sturm, Chris Nowak, Ashley Tate, John Redd, Colin Shepard, Vik Kheterpal, Steven R Steinhubl, Eric J Topol

## Abstract

**Background:** Long COVID affects an estimated 400 million people worldwide, and is associated with low quality of life. Nearly all completed Long COVID clinical trials reported no benefit, and most required participants to travel to study sites. This requirement systematically excludes severely affected patients. Because there are numerous candidate therapeutics with established safety profiles and regulatory approvals for other indications, scaled, efficient evaluation of therapeutics is needed.

**Methods:** We designed and are conducting a double-blind, placebo-controlled, phase two trial of tirzepatide for Long COVID fatigue, using an entirely remote infrastructure. Design elements included electronic consent, identity and diagnosis verification through document upload, cold-chain delivery of an injectable study drug through a central pharmacy, shared decision-making for dose titration, repeated at-home capillary blood collection in a biospecimen subcohort, weekly participant touch points through study application, wrist-worn wearable monitoring, and clinical support. The trial is operating under FDA Investigational New Drug authorization.

**Results:** This trial enrolled 1,058 participants in 73 days, at least double the rate of any other Long COVID trial. Mean baseline metrics include mean Fatigue Severity Scale of 59.3 (standard deviation [SD] 4.9), daily step count of 3,611 (SD 2,706, general population reference mean 7,731), EQ-5D-5L of 0.6 (SD 0.2), and FUNCAP27 4.0 (SD 1.0), which was a more severely affected population than other clinical trials that collected comparable data. Study processes are working as designed. Participants use existing advocacy and support channels to gather and communicate.

**Conclusions:** A direct-to-participant, siteless infrastructure can support a double-blind placebo-controlled trial of an injectable drug at scale, accelerate accrual, and reach severely affected participants who are routinely excluded by site-based designs. Modernizing drug distribution and regulatory pathways is needed to realize the full potential of decentralized infrastructure for drug repurposing clinical trials.

## Introduction

Requiring research and clinical trial participants to travel to a study site creates barriers to participation.^1^ While more research studies and clinical trials have been made available remotely,^2–4^ to our knowledge a placebo-controlled trial of an injectable drug has never been carried out through a fully remote design.

Long COVID affects over 400 million individuals worldwide,^5^ leading to one of the lowest quality of life scores of disease states,^6^ and an estimated $1 trillion annual economic impact.^7^ Half of people with Long COVID develop Myalgic Encephalomyelitis/Chronic Fatigue Syndrome (ME/CFS),^8^ an energy-limiting condition that leaves about 25% of patients housebound, and some bedbound.^9,10^ Long COVID research receives an estimated 14% of the funding it should based on the disability it causes^11^ and while several Long COVID clinical trials have released results, nearly all have reported no benefit.

The combination of severe disease burden and numerous plausible therapeutics suggests drug repurposing is a near-term path to treatments. Effective drug repurposing, especially for an illness as heterogeneous as Long COVID, requires efficiently evaluating many candidates — a problem that the conventional clinical trial practices are poorly suited to solve. Requiring participants to travel to a study site creates barriers to participation, especially for energy-limiting conditions like Long COVID where the barriers exclude the most severely affected patients, resulting in trials that represent a biased subset of the patient population. To our knowledge, all completed and ongoing Long COVID clinical trials except one^2^ have required in-person visits to a trial location.

Here we report on the design and conduct of the Long COVID Treatment Trial – Tirzepatide (LoCITT-T), an entirely remote, double-blind, placebo-controlled trial that enrolled 1,058 participants to evaluate the effect of tirzepatide on Long COVID fatigue. We describe the design choices that made remote conduct feasible for an injectable, refrigerated, Investigational New Drug (IND)-required study drug; the operational performance of the resulting infrastructure; and the lessons that have emerged for future decentralized trials, particularly those evaluating repurposed therapies in post-viral and other energy-limiting conditions.

## Methods

### Trial design and regulatory framework

LoCITT-T is a double-blind, placebo-controlled, parallel-arm, phase two trial of tirzepatide versus matching placebo for Long COVID fatigue, conducted entirely remotely. The trial operates under FDA Investigational New Drug (IND) authorization, was reviewed and approved by the Scripps Institutional Review Board, and was registered on ClinicalTrials.gov (NCT07128082).

Study tasks, including informed consent, surveys, medication administration, and adverse event reporting, are completed through MyDataHelps (MDH), the study application. The staff-facing portion of MDH supports remote review by study nurses and physicians, who communicate with participants by message, phone, or video as needed. Participants may invite a caregiver to assist with any study task and may contact the study team, including nurses, at any time. Study physicians review eligibility (as needed), prescribe study drug, and address clinical questions and issues as they arise.

### Eligibility, consent, and remote identity verification

We utilized centralized infrastructure for all aspects of participant verification and enrollment. Participants were recruited primarily through earned media, social media, webinars, and with a small paid social media campaign focused on engaging Black, Indigenous and People of Color (BIPOC) participants. Before recruitment began, we collected contact information for people who wanted to be notified when we began enrollment for a Long COVID clinical trial; these individuals were notified when enrollment began. Prospective participants reviewed the study website, completed an eligibility survey (Table S1) and uploaded documentation of their Long COVID diagnosis and their identity. Study staff manually reviewed each submission to confirm they met eligibility criteria and that the identity on medical documentation matched their identity verification document, or asked participants to resubmit documents. Next, participants were invited to review and complete the electronic informed consent.

### Study drug delivery and dose titration

Drug distribution is also completed centrally, from one pharmacy directly to participants. Separately, both tirzepatide and placebo are bulk shipped from Eli Lilly and Company to its pharmacy partner, Eversana. Prescription and participant contact information is sent from MDH to Eversana, then Eversana calls participants to arrange shipment to a location of the participant’s choice. Because tirzepatide is temperature-sensitive, deliveries require a signature; participants may designate another individual to receive the package, accommodating those for whom signing for deliveries is not feasible. Each shipment contains four pre-filled syringes, and refill processes begin about two weeks after delivery.

Participants begin on 2.5mg weekly doses, and are given the option to increase their dose by 2.5mg every four weeks with a maximum dose of 15mg; dosing decisions are made jointly by participants and the study clinical team. Participants experiencing side effects are recommended to maintain their dose. Participants who cannot tolerate the starting dose of 2.5mg are given the option to pause dosing or discontinue altogether. This structure was selected to support tolerability in a population where dosing schedules that are standard in other indications often result in intolerable side effects.^10^

Participants were trained on injection technique through video instruction, with the option of a one-on-one call with a study nurse or attending an online webinar. Webinar attendees could ask questions and have them answered during the webinar and then were required to document attendance and confirm understanding of the material.

### Outcomes and assessments

The study’s primary endpoint is the Fatigue Severity Scale, with an eligibility criteria for moderate to severe fatigue (minimum score of 36; full scale ranges from 9 to 63).^12^ This endpoint was selected due to the prominence of fatigue in Long COVID,^13^ and the hypothesis that tirzepatide-mediated reduction in neuroinflammation would attenuate fatigue.^14–16^ The estimand is the difference in Fatigue Severity Scale scores between month 12 and baseline across treatment and control groups for participants that successfully start the study drug, with an interim analysis of FSS at month three to consider early stopping in the case of large effects.

We utilize five additional surveys to enable secondary and exploratory analyses where quarterly surveys are compared to baseline scores (Table 1): 1) changes in overall health, measured by EQ-5D-5L,^17,18^ 2) presence of post-exertional malaise, measured by DSQ-PEM,^19^ 3) functional capacity, measured by FUNCAP27,^20^ 4) Postural Orthostatic Tachycardia Syndrome (POTS), measured by Malmo POTS symptom score (MAPS),^21^ 5) Medication compliance as measured in study-specific weekly check in surveys. Exploratory analyses will include associations between wearable device data (heart rate, HRV, steps, sleep, when available) and changes in symptom burden between time points.^22,23^

**Table 1.** Schedule of events. Months are counted from study drug start date.

| Activity | Always available | Screening | Eligibility | Account creation | Onboarding | Randomization | Just before study drug start | Study drug start | Weekly | Monthly | 3 months | 6 months | 9 months | 12 months |
| --- | --- | --- | --- | --- | --- | --- | --- | --- | --- | --- | --- | --- | --- | --- |
| Screening survey |  | X |  |  |  |  |  |  |  |  |  |  |  |  |
| Identity and diagnosis verification |  |  | X |  |  |  |  |  |  |  |  |  |  |  |
| Informed consent |  |  |  |  | X |  |  |  |  |  |  |  |  |  |
| Baseline demographic survey |  |  |  |  | X |  |  |  |  |  |  |  |  |  |
| Medical history and medication survey |  |  |  |  | X |  |  |  |  |  |  |  |  |  |
| DSQ-COVID |  |  |  |  | X |  |  |  |  |  |  |  |  | X |
| Biosample collection |  |  |  |  |  |  | X |  |  |  |  | X |  | X |
| Medication compliance |  |  |  |  |  |  |  | X | X |  |  |  |  |  |
| Weight check |  |  |  |  | X |  |  | X | X |  |  |  |  |  |
| Monthly refill survey |  |  |  |  |  |  |  | X |  | X |  |  |  |  |
| Fatigue Severity Scale |  |  | X |  | X |  |  | X |  |  | X | X | X | X |
| EQ-5D-5L |  |  |  |  | X |  |  | X |  |  | X | X | X | X |
| MAPS (Malmo POTS) |  |  |  |  | X |  |  | X |  |  | X | X | X | X |
| DSQ-PEM |  |  |  |  | X |  |  | X |  |  | X | X | X | X |
| FUNCAP27 |  |  |  |  | X |  |  | X |  |  | X | X | X | X |
| Adverse event reporting | X |  |  |  |  |  |  |  | X |  |  |  |  |  |

### Statistical analysis

We use multivariate Generalized Estimating Equations for the analysis of our primary endpoint (protocol in supplemental materials). Demographic controls include days since illness onset, POTS status, ME/CFS status, Long-COVID symptom cluster, income status, age, sexual orientation, birthsex, gender, race and/or ethnicity. Supplement 2 provides detailed operationalization (Tables S2; S3).

Prespecified power calculations assume an average FSS of 40.9 points and a standard deviation of 15.8, based on prior work^24^ a range of within-participant correlation rates of 0.3, 0.5 and 0.7, and range of different attrition levels (10% - 30%). Based on our prior Long COVID research, we projected about 13% attrition at the 3-month mark, which provides an MDES (Minimum Detectable Effect Size) of 2.52-2.6 depending on the within-participant correlation rate. At 12-months, we expect attrition to be 29%, which gives an MDES of 2.68-3.06 points.

### Safety monitoring

Adverse event reporting was structured to function without scheduled in-person assessments. Participants may report adverse events through MDH, email, or phone call, at any time. The study team reviews them as soon as possible, including on weekends, and asks participants follow-up questions as needed. Weekly, participants are asked to confirm medication administration, report adverse events, and record weight. Monthly, participants are asked to update their current list of medications, confirm pregnancy status, and complete a suicide risk assessment. Participants who report becoming underweight are contacted by a study nurse to discuss dose adjustment and/or nutritional support.

Severe adverse events are reported to an independent, study-specific Data Safety Monitoring Board (DSMB) within 24 hours of study team awareness, and to the IRB within one week of knowledge of occurrence, including any relevant DSMB response. All unexpected fatal or life-threatening adverse reactions are reported to the FDA within one week of knowledge of their occurrence. This includes instances of suicidal ideation, which are estimated to occur in 21% of people with Long COVID.^25^

### Wearable and biospecimen collection

Once participants enroll and complete baseline surveys, they are eligible to order their study drug, wearable sensor – a Garmin vivosmart 5 based on our prior review of options^26^ – and scale to enable weekly weight reporting. These devices are available to participants at no cost to them and shipped in a teal study-branded box, the color chosen by the Long COVID community to represent hope and support.^27^

Blood samples are collected from a 50-participant subcohort at baseline, 6 months, and 12 months using at-home capillary blood draw devices. Samples are analyzed for clinical labs (HbA1c, CRP, cortisol) and proteomics (Olink Reveal; Infinity Bio HuSIGHT and VirSIGHT). Changes over time are evaluated across study arms.

## Results

### Recruitment and Participation

LoCITT-T enrolled 1,058 participants over 73 days (Figure 1). The eligibility survey opened on October 29, 2025 and closed on December 2, 2025, a 35-day window during which prospective participants completed screening. Enrollment completion continued through January 5, 2026 as participants completed identity verification, diagnosis documentation, and informed consent. The largest single-day recruitment volume (n=173 screened in) coincided with a public webinar in which the study team answered questions.^28^ The pace was at least 2.5 times as fast as all other evaluable Long COVID trials (Figure 2, Table S5). This was enabled by a combination of strategies. First, people with Long COVID, caregivers, and allies (subsequently referred to as the Long COVID community) have convened online since the early days of the COVID pandemic.^29^ Support and advocacy networks helped trial information spread quickly, along with emerging anecdotal reports that GLP-1 receptor agonists helped with Long COVID symptom management.^30^ Second, 533 individuals signed up to be notified when enrollment began. Third, because the trial was available across the United States (except for Hawaii, due to shipping constraints), more individuals were eligible than trials that require visits to specific locations.

**Figure 1.**
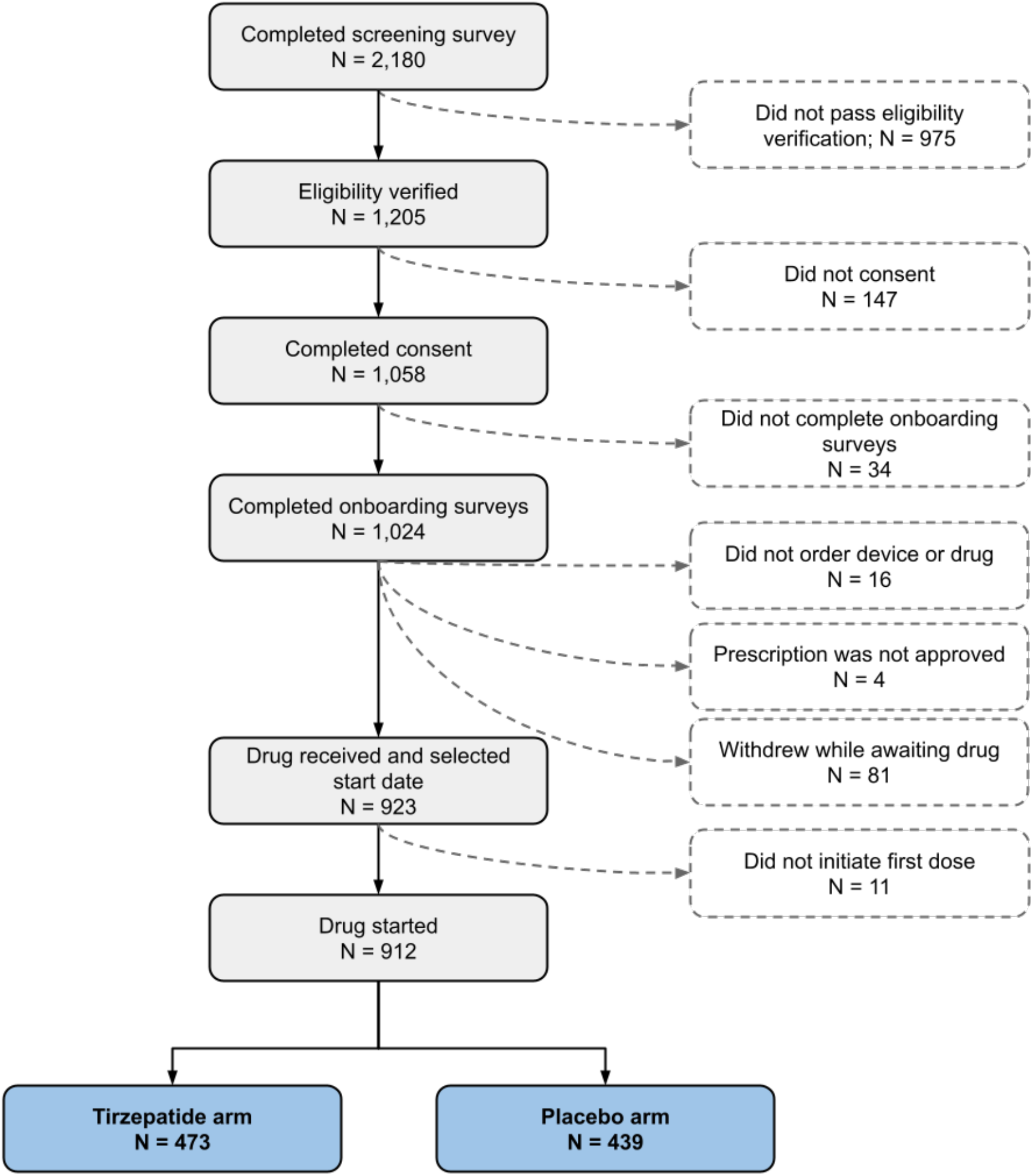
Diagram displaying number of individuals at each stage from screening through initiating drug.

**Figure 2.**
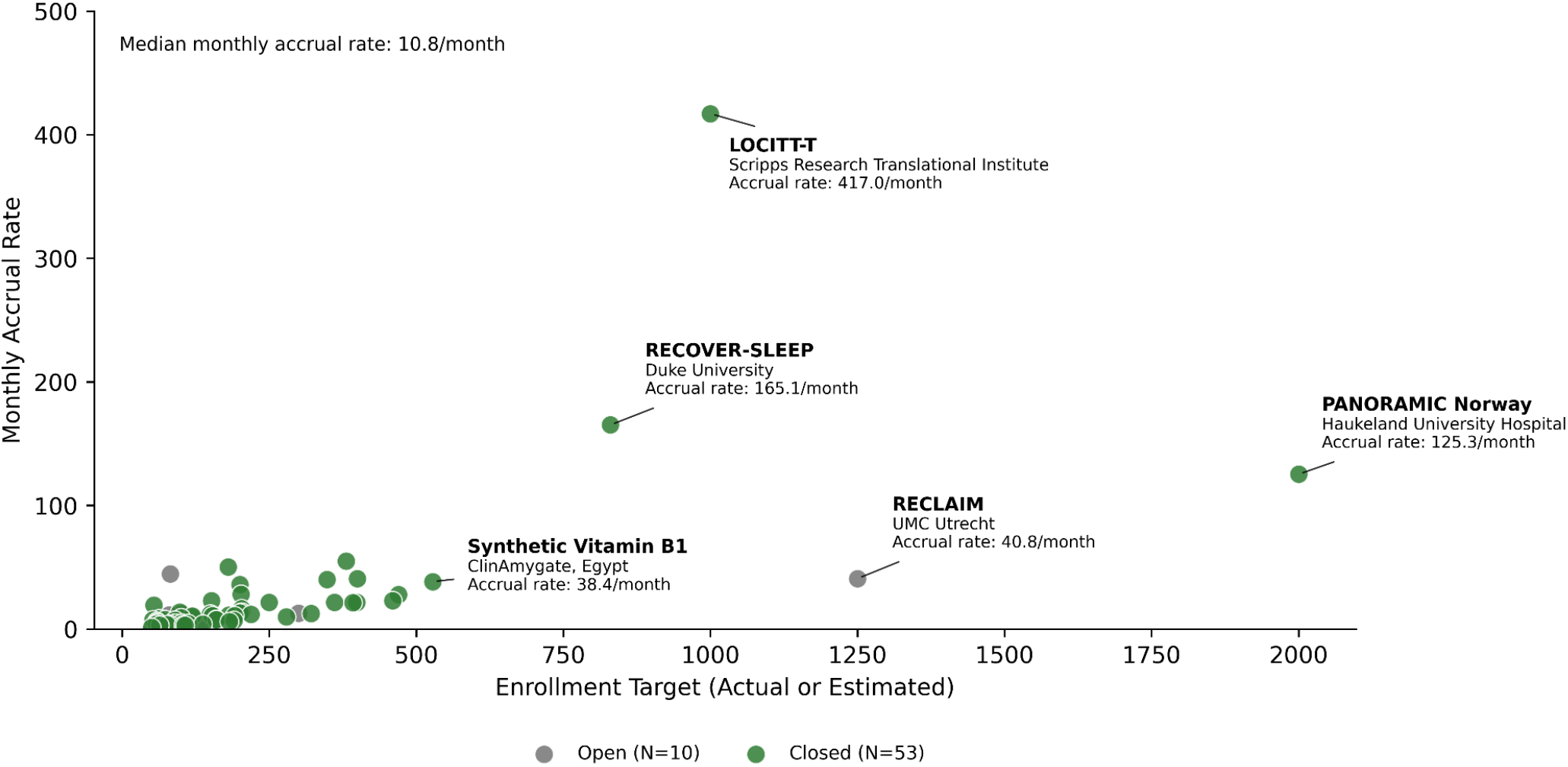
Enrollment timelines of Long COVID trials (n=63) based on data from clinicaltrials.gov.

**Figure 3.**
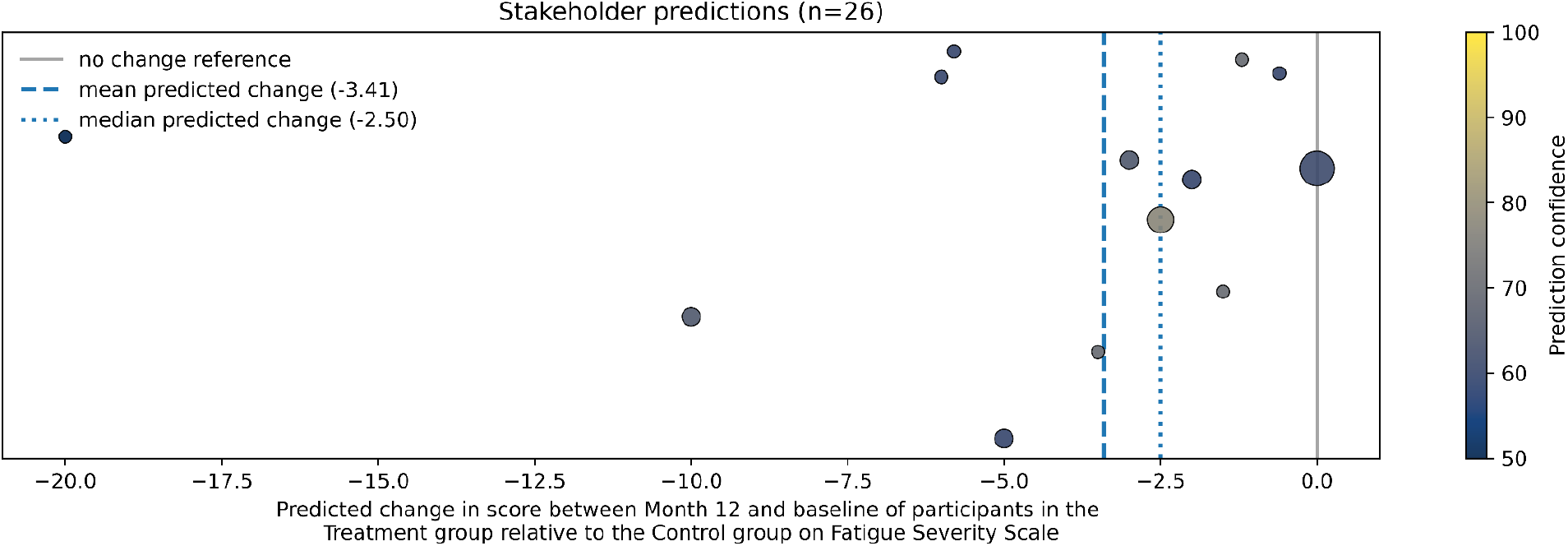
Results of request to predict trial outcome sent to key stakeholders including: study staff, people with Long COVID, Long COVID researchers, Long COVID health care providers, and trial funders. Confidence of 50% represents a coin flip and 100% represents certainty. Note that 3 respondents stated that they expected “no difference in fatigue between the drug and treatment group” but then subsequently declared non-zero effect size estimates (values 1, 2.5 and 3). These were re-coded to zero for the analysis.

Assignment to tirzepatide and placebo was balanced (473 tirzepatide; 439 placebo; p=0.22). Of those who completed baseline surveys, 112 participants (10.9%) did not initiate study drug. Commonly cited reasons were a change in medical circumstance (21.9%), medication shipping delays (9.6%, median delay: 50 days), and accessing tirzepatide outside the trial (4.0%). Enrollment continued until 1,000 active participants ordered study device kits.

### Study progression and compliance

As of July 2026, 911 participants (99.9%) started the study drug at least 3 months ago. For those continuing in the study, 95.5% completed all 5 quarterly check-in surveys; 96.4% completed FSS. Drug administration rate is calculated as the percentage of participants that report having administered the drug each week, divided over the number of weeks that have passed since starting the drug; rates average 91.7% for the cohort (IQR: 90.9% - 100%). When a participant agrees with the study team to skip a dose it is counted as a week without drug administration.

### Patient-centric design

Before beginning recruitment, the trial design was evaluated using a Patient-Led Research scorecard^31^ completed independently by people with Long COVID and by study staff without Long COVID (mean 4.58 out of 5, standard deviation 0.29, Figure S1). In support of conducting a patient-centric trial, we drafted communication guidelines to support both cohort-wide participant communications and individual interactions (Table S4).

### Baseline data and comparison to other trials

Baseline characteristics indicated that the trial reached a severely affected and geographically dispersed cohort (Table 2). The most common FSS score was the highest possible score, indicating the worst measurable fatigue; therefore, participants could have changes in fatigue that are not detectable. The average EQ-5D-5L score in our cohort indicates quality of life as low or lower than other cohorts.^6,32–34^ We compared baseline data to several other trials and our cohort had more severe symptoms, with the other remote trial having the closest severity (Table 2).^35–39^ We examined the number of prescription medications that participants reported taking at baseline; the median was 2 (IQR 1-6), and 32.5% (n=296) met the definition of polypharmacy with at least five prescription medications.^40^

**Table 2.** Baseline metrics across the LoCITT-T cohort (n = 912) and from other trials that collected a data type in common with LoCITT-T, and either had at least 200 participants, was a remote trial (PAX-LC) or collected a more rare data type (STOP-PASC wearable substudy).

| Metric | Mean | Standard deviation | Score indicating best measurable health | Score indicating worst measurable health |
| --- | --- | --- | --- | --- |
| FSS | 59.3 | 4.9 | 9, however eligibility required score of at least 36 | 63 |
| EQ-5D-5L | 0.6 | 0.2 | 1 | -1.29 |
| EQ-5D-5L level sum score | 12.3 | 3.5 | 5 | 25 |
| FUNCAP27 | 4.0 | 1.0 | 6 | 0 |
| DSQ-PEM (composite score) | 64.5 | 21.4 | 0 | 100 |
| DSQ-PEM (PEM incidence based on binary score) | 0.9 | 0.3 | 0 | 1 |
| Malmo POTS | 54.8 | 23.2 | 0 | 120 |
| Daily step count* | 3,611 | 2,706 | General population average, 7,731 | 0 |
| * Daily step count at baseline reflects the average number of steps recorded over the first 7 days after the participant has started wearing the device (ie., first non-zero heart rate or step count value). |  |  |  |  |

| Metric | 10th percentile | 25th percentile | 50th percentile | 75th percentile | 90th percentile |
| --- | --- | --- | --- | --- | --- |
| FSS | 53.0 | 57.0 | 61.0 | 63.0 | 63.0 |
| EQ-5D-5L | 0.3 | 0.5 | 0.7 | 0.8 | 0.9 |
| EQ-5D-5L level sum scores | 8.0 | 10.0 | 12.0 | 15.0 | 17.0 |
| FUNCAP27 | 2.6 | 3.3 | 4.1 | 4.8 | 5.1 |
| DSQ-PEM (composite score) | 35.0 | 50.0 | 67.5 | 80.0 | 92.5 |
| DSQ-PEM (PEM incidence based on binary score) | 1 | 1 | 1 | 1 | 1 |
| Malmo POTS | 24.1 | 37.0 | 54.0 | 72.0 | 86.0 |
| Daily step count* | 785 | 1,664 | 3,136 | 4,901 | 6,930 |

| Metric | Value | Trial | Study N | % difference |
| --- | --- | --- | --- | --- |
| FSS median | 52.7 | REVIVE<sup>36</sup> | 399 | 13.7 |
| FSS mean | 46.1 | Prospekta<sup>38</sup> | 670 | 22.1 |
| EQ-5D-5L level sum score | 11.4 | PAX-LC | 93 | 8.0 |
| Daily step count | 4,311 | STOP-PASC wearable substudy<sup>39</sup> | 50 | 19.3 |
| Daily step count | 5,593 | RECOVER digital health platform<sup>22</sup> | 498 | 54.8 |

We also compare the trial cost to other clinical trials (Table 3). While trial costs can vary widely based on the design, LoCITT-T per participant costs are less than one tenth those of NIH-funded Long COVID trials.^41,42^

**Table 3.**
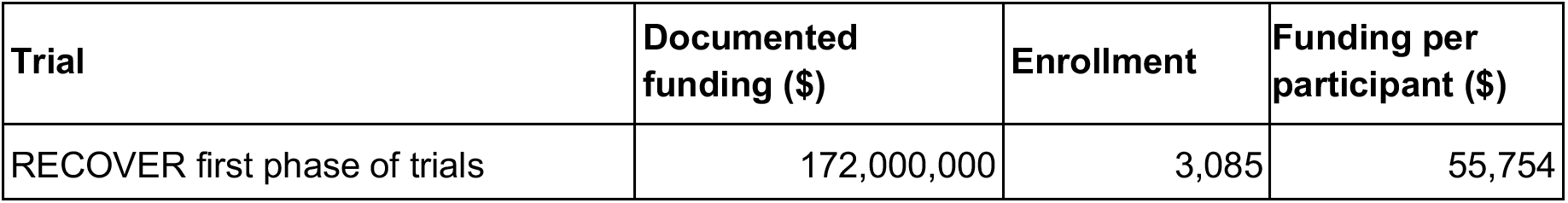
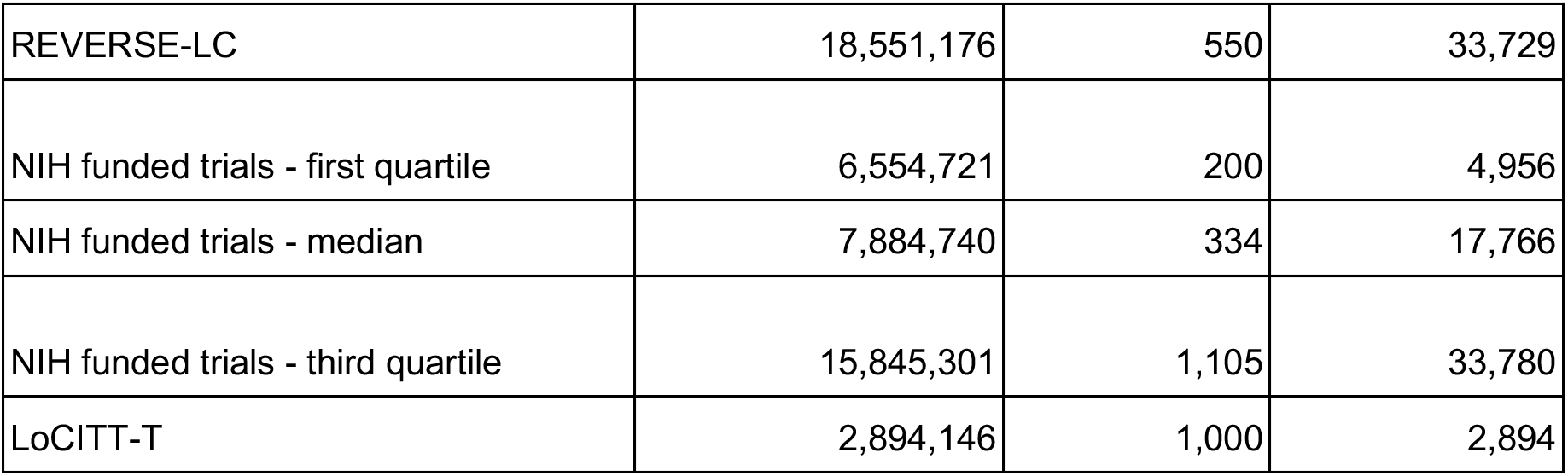
Total funding and funding per participant across trials with publicly available data compared to LoCITT-T. Clinical trials were searched on NIH Reporter for all trials funded in fiscal years 2020 to 2026, and enrollment numbers were cross-referenced with clinicaltrials.gov. Only trials with both data sources were included. For trials that are ongoing, it is likely that costs are undercounted because funding for later fiscal years has not yet been included. Even with that under-reporting LoCITT-T is among those with the lowest cost per participant. Data on all included trials is in Supplement 6.

### Long COVID Community

Participants in this trial have connected with each other, forming private online groups on multiple platforms. For example a participant-organized Facebook group has 282 members who moderators stated were vetted to ensure they were trial participants. While the study team does not participate in these groups, we have been made aware that their content includes sharing study events such as webinars, experiences with study drug, misinformation, and peer support. We understand the natural tendency to convene given the shared experience and that there is substantial publicly available information about how people with Long COVID respond to tirzepatide. We have requested that participants bring all medical questions to the study team and refrain from active efforts to unblind themselves and others.

Many participants have expressed concerns about being in the placebo group and questioned the value of their continued participation; we have reinforced that the placebo group is essential to accurately assess the effect of tirzepatide. We were made aware of one participant that sent their study drug for testing to determine whether it contained tirzepatide and withdrew upon learning they were in the placebo arm. In the future, we will update our consent form to clarify that attempting unblinding is not permitted. While we explored conducting an open label extension, it was not feasible for this trial; we strongly recommend that trials include an open label extension both to give equitable access to the drug across trial participants and to collect additional data about the drug’s effects in an unblinded setting.

Due to information-sharing within the Long COVID community, participants were also well informed about tirzepatide dosing and many requested the option for micro-dosing, e.g. taking one tenth of the standard starting dose. Based on the use of pre-filled syringes, which contain a non-preserved formulation without graduated markings, any dose less than the complete injection of prefilled syringe (with doses as low as 2.5mg), including micro-dosing, could not be given accurately and safely and was not supported. When beginning medications, people with Long COVID and/or ME/CFS often benefit from the strategy of “start low, and go slow” and we recommend that future drug repurposing trials consider starting with lower dosing and allow for slower titration schedules than is used for other indications.^10,43^

### Safety Monitoring

The trial’s DSMB determined that all participants reporting suicidal ideation, who must be withdrawn from the study due to no longer meeting the eligibility criterion, are unblinded so that if they were taking tirzepatide, this can be considered in their future healthcare. This information was of particular interest to the DSMB because in January 2026, FDA requested the removal of suicidal ideation and behavior as a risk associated with GLP-1 receptor agonists with weight management indications.^44^ Additionally, the risk of suicidal ideation may be higher in people with Long COVID.^25^

Participants were provided the opportunity to report adverse events in an open text field, an intentional choice to avoid biasing toward suggested side effects. One challenge this introduced was differentiating between symptoms that are new and potentially study-related versus ongoing. Due to the high and dynamic baseline symptom burden, adverse events are sometimes only identified by the participant as distinct from baseline when there is a significant change from their baseline. Additionally, the open text format enabled detailed narrative reporting. Across 5,797 adverse event reports, length varied from 1 to 575 words (mean 21, SD 35, median 9, IQR 4-24). This reflects commendable participant engagement, and requires substantial effort to systematically identify adverse events.

### Expected trial results

Prior to starting data analysis, we asked key stakeholders to share what they expected the trial’s results to be and how confident they were. These predictions help us anchor eventual results and increase accountability and learning^45^. Based on 26 responses, the median predicted change is a 2.50 point reduction in FSS in the tirzepatide group relative to the placebo group (Figure S1). While most predictions indicated that tirzepatide is expected to decrease fatigue, seven individuals predicted no effect, and no stakeholder predicted that tirzepatide would increase fatigue. The median confidence of 65% was relatively low. Most stakeholders predict a smaller positive effect of tirzepatide than what we are powered to detect.

### Operations

Initial study drug inventory was available later than expected, which led to a compressed timeframe for most participants to start dosing. While we forecasted that dosing titration would be slower than in other trials, study drug availability was not initially aligned with those projections, leading 9.6% of participants to wait 3 months to receive their first dose and 38.9% of participants to have at least one shipping delay of at least 11 days after starting medication. Subsequently, we worked to more proactively track and project inventory. Another challenge is that the pharmacy required an initial synchronous phone call, which some participants find burdensome. Later shipping could be coordinated by text, which is often more accessible. Signature for deliveries sometimes presented a logistical barrier. When using overnight shipping in general, delays and mistakes affect 5-10% of shipments,^46,47^ and we recommend planning for 10 to 15% more inventory than is required for any given trial.

We initially requested a pre-IND meeting with FDA, expecting that the trial would be exempt from IND requirements because tirzepatide is FDA approved, the trial data would not be used as the sole basis for requesting a label expansion, and we expect no substantive increased risk in the trial population versus the populations tirzepatide is approved in.^48^ However, we were required to obtain an IND. This resulted in a delayed timeline, as FDA is required to respond to a pre-IND meeting request within 60 days^49^ and required to respond to an IND submission within 30 days,^48^ and we completed both.

## Discussion

LoCITT-T establishes the feasibility of conducting a large double-blind, randomized controlled trial of an injectable drug using completely decentralized infrastructure. This process and infrastructure are scalable and replicable, and can be leveraged to rapidly conduct additional trials, with flexibility to extend to designs such as platform trials, and other modes of drug administration. Future work will include expansions to incorporate elements of in-person interactions, such as intravenously-administered drugs and phlebotomist visits.

In Long COVID and other energy-limiting conditions, enabling participation from home is crucial for including those who are most severely affected by the illness. Based on baseline data we recruited a more severe cohort than other Long COVID clinical trials. Further, on a per participant basis, LoCITT-T is less costly than nearly all other evaluable trials. While each trial has its own complexity and duration, when remote participation is offered it can reduce participant effort and financial burden. LoCITT-T infrastructure enabled accrual at 2.5 times the pace of other trials. Operational challenges arose at the interfaces with partners whose workflows were accustomed to a site-based trial model: drug supply and shipping, and regulatory pathways. Conducting efficient decentralized trials requires all parties to adopt workflows that center on participants and enable timely action.

Two noteworthy participant behaviors arose in this trial that may be relevant in subsequent trials. First, people with Long COVID have built active online communities that exist independently of any trial.^29^ This was beneficial to trial enrollment and potentially detrimental when participants connect to compare trial experiences, including one recorded case of independent testing of study drug. Second, since the study drug is FDA approved and available through prescription or off-label channels, some participants viewed suspected placebo assignment as carrying an opportunity cost that may not be present in trials of novel therapeutics, although the drug is unaffordable for many.^50^ Some participants withdrew when they were able to access tirzepatide outside the trial, and others questioned the value of continued participation if they suspected placebo assignment. Future trial design should anticipate these challenges by including consent language stating that attempts to determine treatment assignment are not allowed because they compromise trial integrity, and communication strategies with a trusted channel for medical questions. Further, open-label extensions should be considered both as an equity measure and as a retention mechanism that addresses the marketplace-alternative pressure on placebo retention.

LoCITT-T demonstrates that fully decentralized infrastructure can support a large, rigorous, double-blind, placebo-controlled trial of an injectable drug, with rapid recruitment, in a patient population that some trials have struggled to enroll. By centralizing infrastructure rather than replicating it across physical locations, a cost-effective model was deployed. These strategies could be used to trial multiple repurposed drugs in parallel, enabling more rapid advancement. Additional improvements are largely logistical, proactively managing drug supply and delivery, and streamlining regulatory pathways. The processes described herein offer a basis for the clinical trials to evaluate candidate therapies at a more efficient pace and scale.

## Data Availability

Data are part of an ongoing, blinded clinical trial and will be made available with the publication of trial results.

## Acknowledgements

We are grateful to all trial participants for their efforts.

## Supplemental Materials

For the manuscript entitled:

Insights from a double-blind, randomized, direct-to-participant intervention trial for Long COVID

### 1. Eligibility criteria

**Table S1.** LoCITT-T inclusion and exclusion criteria.

| Category | Criterion |
| --- | --- |
| **Inclusion Criteria** |  |
| **Age** | 18 years of age or older |
| **Residency** | Living in the United States |
| **Language** | Able to read and understand English |
| **Study Participation** | Willing and able to participate in study interventions and activities, including: access to an internet-connected device; completion of informed consent; surveys; medication schedule adherence; adverse event reporting; weight reporting; use of a wearable activity tracker; and completion of at-home blood collections, if selected |
| **Long COVID Diagnosis** | Meets the NASEM definition of Long COVID: an infection-associated chronic condition occurring after SARS-CoV-2 infection, present for at least 3 months as a continuous, relapsing and remitting, or progressive disease state affecting one or more organ systems<sup>1</sup> |
| **Identity Verification** | Ability to verify identity |
| **Diagnosis Verification** | Ability to verify Long COVID diagnosis |
| **Concurrent Treatment Disclosure** | Agreement to notify the study team if any other Long COVID treatments are initiated during enrollment |
| **Fatigue Assessment** | Completion of the Fatigue Severity Scale with a minimum qualifying score of 36 |
| **Exclusion Criteria** |  |
| **Vulnerable Populations** | Members of certain vulnerable populations, including prisoners, minors, fetuses, and institutionalized individuals |
| **Pregnancy** | Pregnant women, due to tirzepatide's unknown risks to the fetus |
| **Thyroid Malignancy** | Personal or family history of medullary thyroid carcinoma |
| **Gastrointestinal Disease** | History of severe gastrointestinal disease |
| **Gastroparesis** | Diagnosis of gastroparesis |
| **Renal Failure** | Worsening or chronic renal failure |
| **Pancreatitis** | History of pancreatitis |
| **Endocrine Disorder** | Multiple Endocrine Neoplasia syndrome type 2 (MEN2) |
| **Drug Hypersensitivity** | Known serious hypersensitivity to tirzepatide |
| **Current GLP-1 Use** | Current use of tirzepatide or another GLP-1 receptor agonist |
| **Medication Contraindications** | Medication contraindications to tirzepatide, as evaluated by Scripps physician when needed |
| **Suicidality** | History of suicidal attempts and/or active suicidal ideation |
| **Low Body Weight** | Body mass index below 18.5 kg/m<sup>2</sup> |
| **Planned Surgery** | Plans to undergo elective surgery or procedures requiring general anesthesia or deep sedation within the subsequent 12 months |
| **Pre-existing Symptoms** | Fatigue and/or cognitive impairment predating SARS-CoV-2 infection |

Abbreviations: GLP-1, glucagon-like peptide-1; MEN2, multiple endocrine neoplasia type 2; NASEM, National Academies of Sciences, Engineering, and Medicine; SARS-CoV-2, severe acute respiratory syndrome coronavirus 2.

### 2. Operationalization of outcomes and control variables

#### Outcome measures

**Table S2:** Data sources and operationalization of each outcome variable.

| Measure | Data source | Operationalization |
| --- | --- | --- |
| Fatigue Severity Scale (FSS) | Fatigue Severity Scale survey, all questions<br>*During the past week, I have found that:*<br>1. My motivation is lower when I am fatigued.<br>2. Exercise brings on my fatigue.<br>3. I am easily fatigued.<br>4. Fatigue interferes with my physical functioning.<br>5. Fatigue causes frequent problems for me.<br>6. My fatigue prevents sustained physical functioning.<br>7. Fatigue interferes with carrying out certain duties and responsibilities.<br>8. Fatigue is among my three most disabling symptoms.<br>9. Fatigue interferes with my work, family, or social life.<br>*Response options: Single punch scale per item: 1-7 (1: Strong disagreement; 7: Strong agreement)* | Scores as summed across all items (range: 9 - 63). |
| Quality of Life (EQ-5D-5L) | EQ-5D-5L survey, all questions<br>*Please select one answer that best describes your health TODAY:*<br>**MOBILITY**<br>• I have no problems in walking about (0)<br>• I have slight problems in walking about (0.058)<br>• I have moderate problems in walking about (0.076)<br>• I have severe problems in walking about (0.207)<br>• I am unable to walk about (0.274)<br>**SELF-CARE**<br>• I have no problems washing or dressing myself (0)<br>• I have slight problems washing or dressing myself (0.050)<br>• I have moderate problems washing or dressing myself (0.080)<br>• I have severe problems washing or dressing myself (0.164)<br>• I am unable to wash or dress myself (0.203)<br>**USUAL ACTIVITIES** (e.g. work, study, housework, family or leisure activities)<br>• I have no problems doing my usual activities (0)<br>• I have slight problems doing my usual activities (0.050)<br>• I have moderate problems doing my usual activities (0.063)<br>• I have severe problems doing my usual activities (0.162)<br>• I am unable to do my usual activities (0.184)<br>**PAIN / DISCOMFORT**<br>• I have no pain or discomfort (0)<br>• I have slight pain or discomfort (0.063)<br>• I have moderate pain or discomfort (0.084)<br>• I have severe pain or discomfort (0.276)<br>• I have extreme pain or discomfort (0.335)<br>**ANXIETY / DEPRESSION**<br>• I am not anxious or depressed (0)<br>• I am slightly anxious or depressed (0.078) | Each response is assigned a value based on a severity weight<sup>2</sup> that is shown in brackets next to each answer option (this is not visible to participants). The values of all 5 questions are summed, then subtracted from the constant 1 to arrive at their final value. |
|  | • *I am moderately anxious or depressed (0.104)*<br>• *I am severely anxious or depressed (0.285)*<br>• *I am extremely anxious or depressed (0.289)*<br>*Response options: single punch per item* |  |
| Malmo-POTS | MAPS survey, all questions<br>1. *Dizziness in upright position or while standing up*<br>2. *Dizziness, feeling that you are going to faint*<br>3. *Palpitations, high pulse, or feeling heart beating irregularly*<br>4. *Difficult breathing/dyspnoea, both at effort and rest*<br>5. *Chest pain*<br>6. *Headache*<br>7. *Concentration difficulties and/or problems with thinking*<br>8. *Muscle pain*<br>9. *Nausea*<br>10. *Gastrointestinal problems (stomach-ache, diarrhea, constipation)*<br>11. *Abnormal tiredness that persists after rest*<br>12. *Insomnia*<br>*Response options: Single punch scale per item: 0-10 (0: No symptoms; 10: Pronounced symptoms)* | Scores as summed across all items (range: 0 - 120). |
| PEM incidence | DSQ-PEM survey, all questions<br>*Throughout the past 3 months:*<br>*How often have you experienced a dead, heavy feeling after starting to exercise?*<br>*Response options (single punch):*<br>• *0: none of the time*<br>• *1: a little of the time*<br>• *2: about half the time*<br>• *3: most of the time*<br>• *4: all of the time* | Binary variable with a value of 1 if the participant responds with a 2 or higher on both the frequency and severity for any given symptom in the survey, 0 otherwise |
| DSQ-PEM composite score | *How much has a dead, heavy feeling after starting to exercise bothered you?*<br>*Response options (single punch):*<br>• *1: Mild*<br>• *2: Moderate*<br>• *3: Severe*<br>• *4: Very Severe*<br>*How often have you experienced next day soreness or fatigue after non-strenuous, everyday activities?*<br>*Response options (single punch):*<br>• *0: none of the time*<br>• *1: a little of the time*<br>• *2: about half the time*<br>• *3: most of the time*<br>• *4: all of the time*<br>*How much has next day soreness or fatigue after non-strenuous, everyday activities bothered you?*<br>*Response options (single punch):*<br>• *1: Mild*<br>• *2: Moderate*<br>• *3: Severe* | Average score across all items, then multiply by 25 (range: 0 - 100). |
|  | • 4: Very Severe<br>*How often have you experienced being mentally tired after the slightest effort?*<br>*Response options (single punch):*<br>• 0: none of the time<br>• 1: a little of the time<br>• 2: about half the time<br>• 3: most of the time<br>• 4: all of the time<br>*How much has being mentally tired after the slightest effort bothered you?*<br>*Response options (single punch):*<br>• 1: Mild<br>• 2: Moderate<br>• 3: Severe<br>• 4: Very Severe<br>*How often have you experienced minimum exercise makes you physically tired?*<br>*Response options (single punch):*<br>• 0: none of the time<br>• 1: a little of the time<br>• 2: about half the time<br>• 3: most of the time<br>• 4: all of the time<br>*How much has minimum exercise that makes you physically tired bothered you?*<br>*Response options (single punch):*<br>• 1: Mild<br>• 2: Moderate<br>• 3: Severe<br>• 4: Very Severe<br>*How often have you experienced feeling physically drained or sick after mild activity?*<br>*Response options (single punch):*<br>• 0: none of the time<br>• 1: a little of the time<br>• 2: about half the time<br>• 3: most of the time<br>• 4: all of the time<br>*How much has physically drained or sick after mild activity bothered you?*<br>*Response options (single punch):*<br>• 1: Mild<br>• 2: Moderate<br>• 3: Severe<br>• 4: Very Severe |  |
| Functional Capacity (FUNCAP) | FUNCAP survey, all questions<br>*What are the consequences for you if you perform the activities described below?*<br>*To what extent does this affect how much else you do?* | Average score across all items (range: 0 - 6). |
|  | *A. Personal hygiene / basic functions*<br>*1. Using the toilet (not bedpan or bedside commode)*<br>*2. Showering standing up*<br>*3. Getting dressed in regular clothes*<br>*B. Walking – moving around*<br>*4. Walking a short distance indoors, from one room to another*<br>*5. Walking between approx. 100 m and 1 km on level ground (length of 1 to 10 football fields)*<br>*6. Physical activity with increased heart rate, for approx. 15 min*<br>*C. Being upright*<br>*7. Sitting in bed for approx. ½ hour*<br>*8. Sitting in an upright chair (dining chair) with feet on floor for approx. 2 hours*<br>*9. Standing up for approx. 5 minutes, e.g. while queuing or while cooking*<br>*D. Activities in the home*<br>*10. Heavier housework (washing floors, vacuuming etc.) for approx. 1/2 hour continuously*<br>*11. Cooking a complicated meal from scratch, approx. 1 hour of preparation*<br>*E. Communication*<br>*12. Having a conversation for approx. 5 minutes*<br>*13. Participating in a conversation with three people for approx. 1/2 hour*<br>*14. Participating in a dinner party, party or family event*<br>*F. Activities outside your home*<br>*15. Stepping right outside your home*<br>*16. Going to a shop for groceries*<br>*17. Using public transport (bus or train)*<br>*18. Participating in organized leisure activities such as classes, sports etc.*<br>*G. Reactions to light and sound*<br>*19. Staying in a room with normal lighting, without sunglasses, for approx. 1 hour*<br>*20. Staying outdoors in daylight without sunglasses for approx. 2 hours*<br>*21. Staying in a noisy environment, (shopping mall, café or open plan office) for approx. 1 hour*<br>*H. Concentration*<br>*22. Reading a short text, such as a mobile phone text message*<br>*23. Reading and understanding a non-fiction text, such as an official document one A4 page long*<br>*24. Using social media to stay in touch with others*<br>*25. Focusing on a task for approx. 10 minutes continuously*<br>*26. Focusing on a task for approx. 2 hours continuously*<br>*27. Managing a full working day (non-physical work such as office work, classes or lectures)*<br>*Response options: Single punch scale per item: 0-6, where:*<br>*• 0: I cannot do this*<br>*• 1: My capacity will be severely reduced for at least three days*<br>*• 2: I can do little else on the same day and for one to two days afterwards* |  |
|  | • 3: I can do little else on the same day<br>• 4: I must limit other activities on the same day<br>• 5: This rarely affects other activities<br>• 6: Unproblematic – does not affect other activities |  |

#### Control variables

**Table S3:**
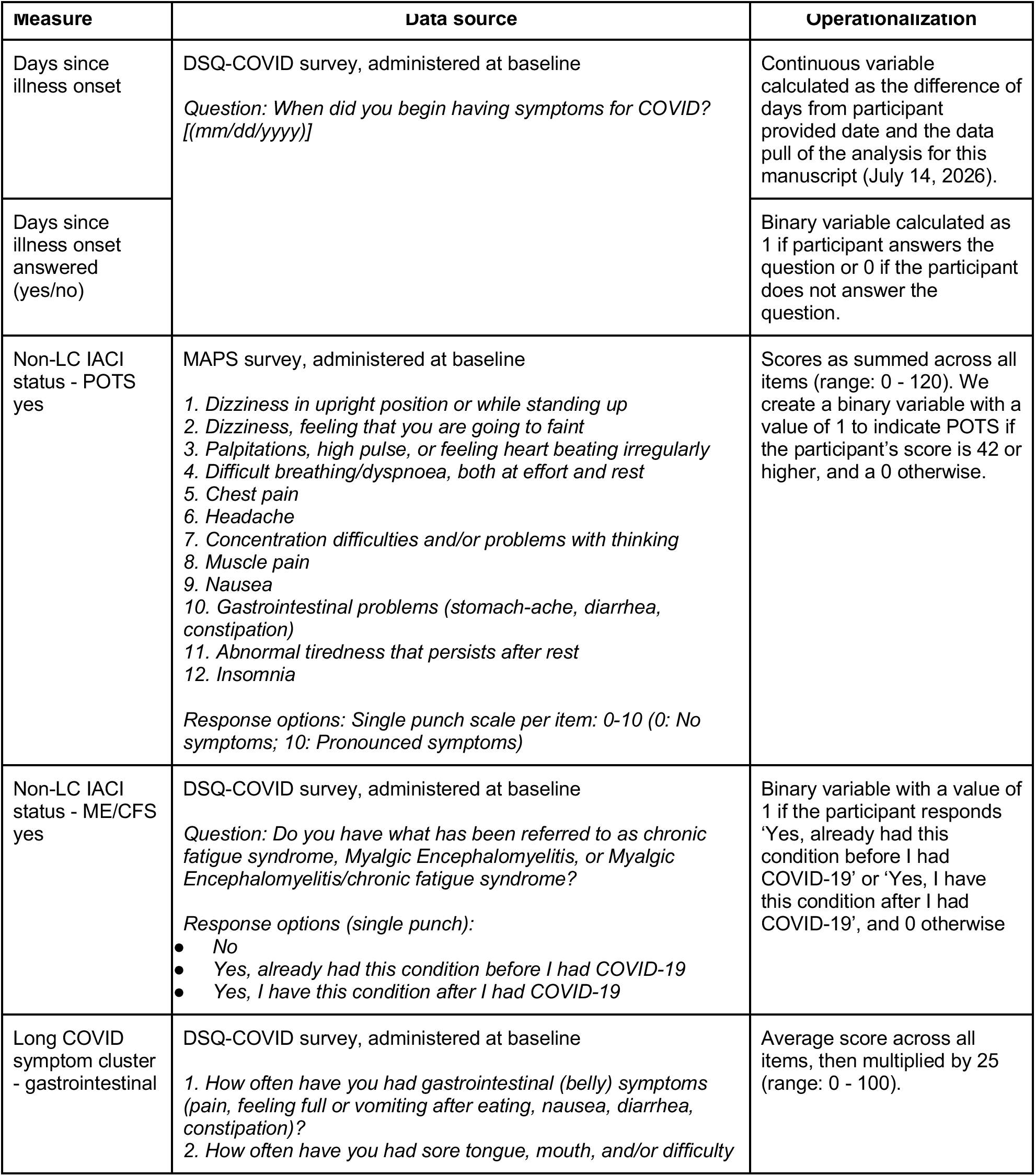
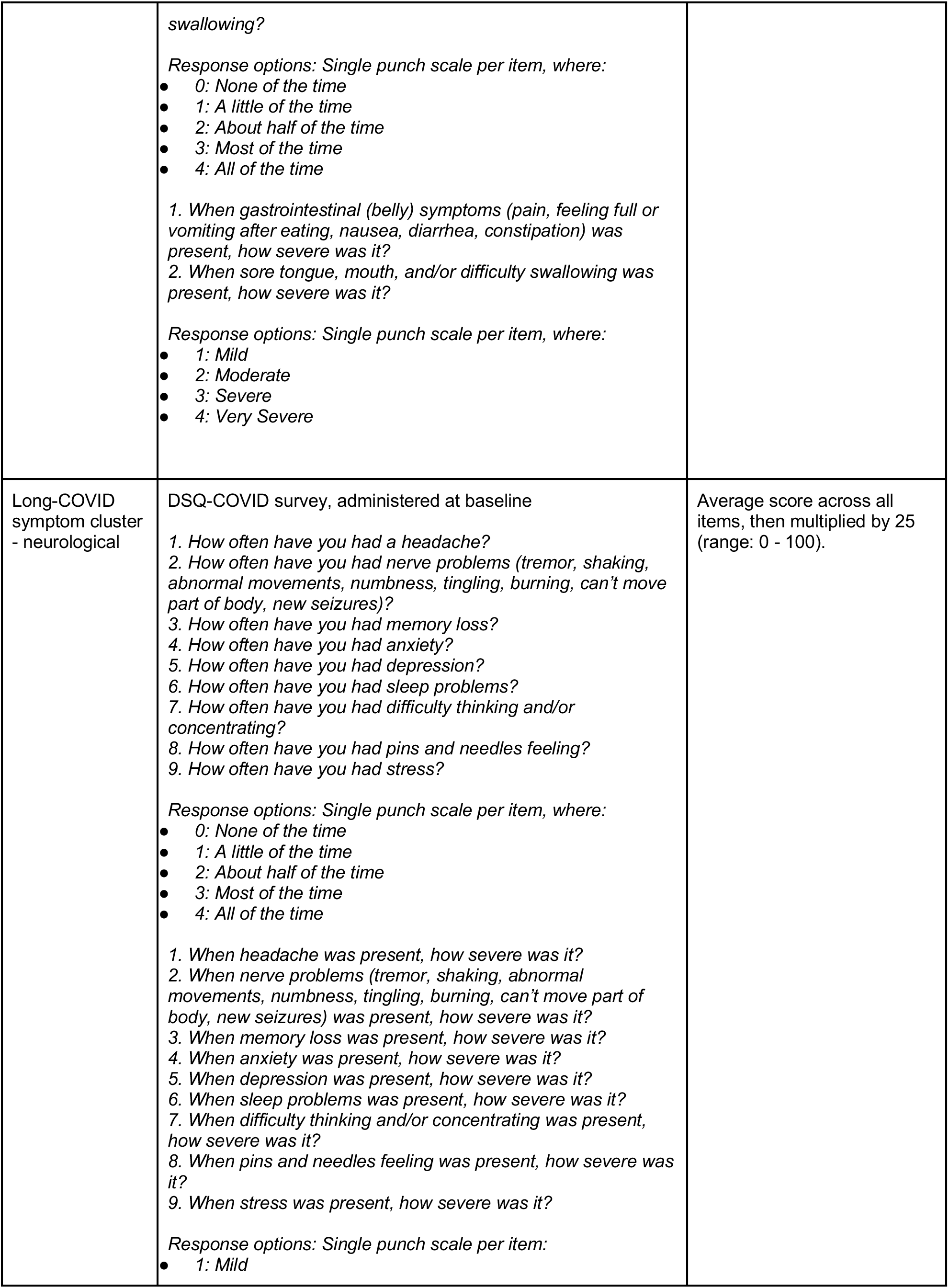
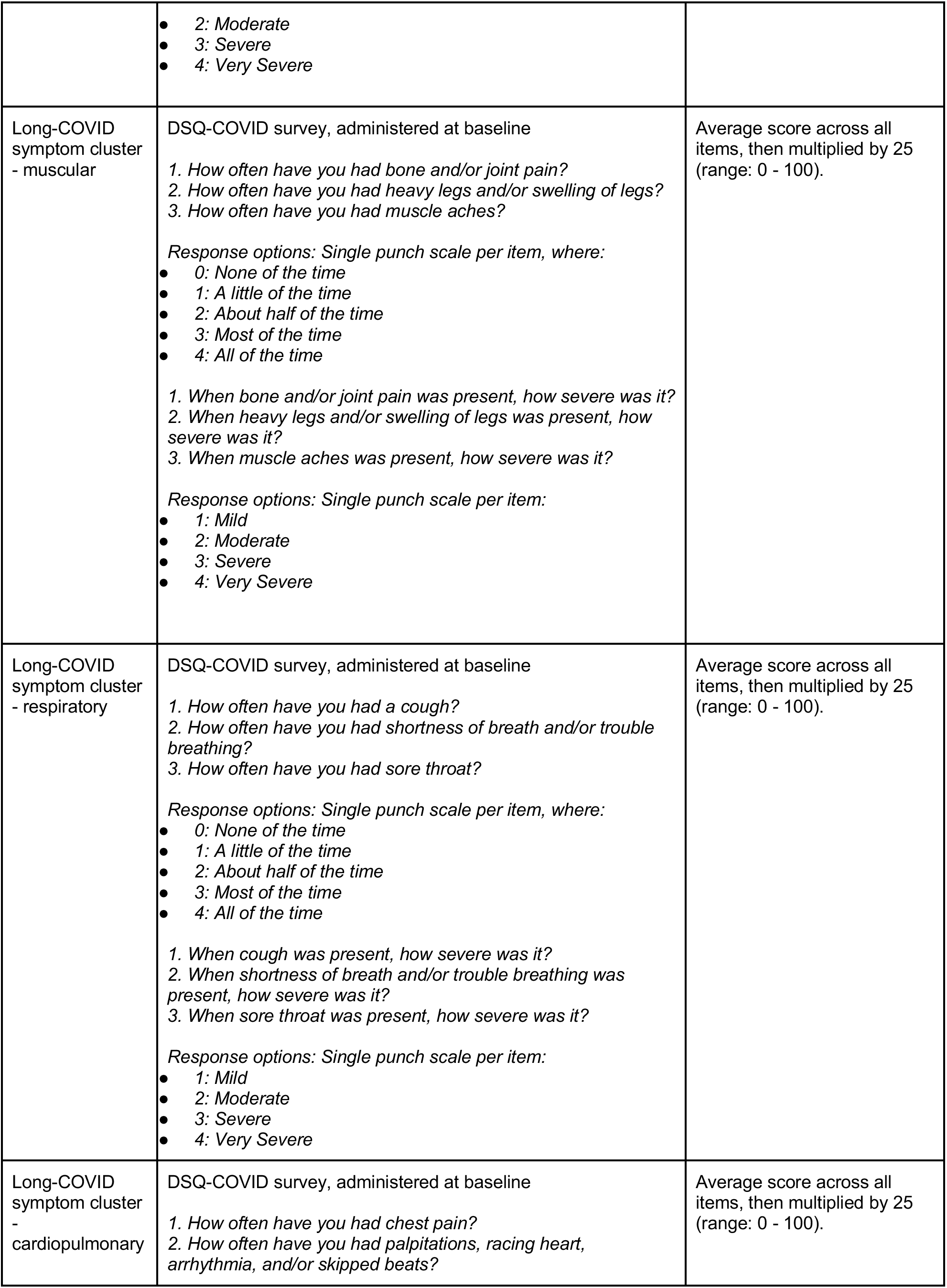
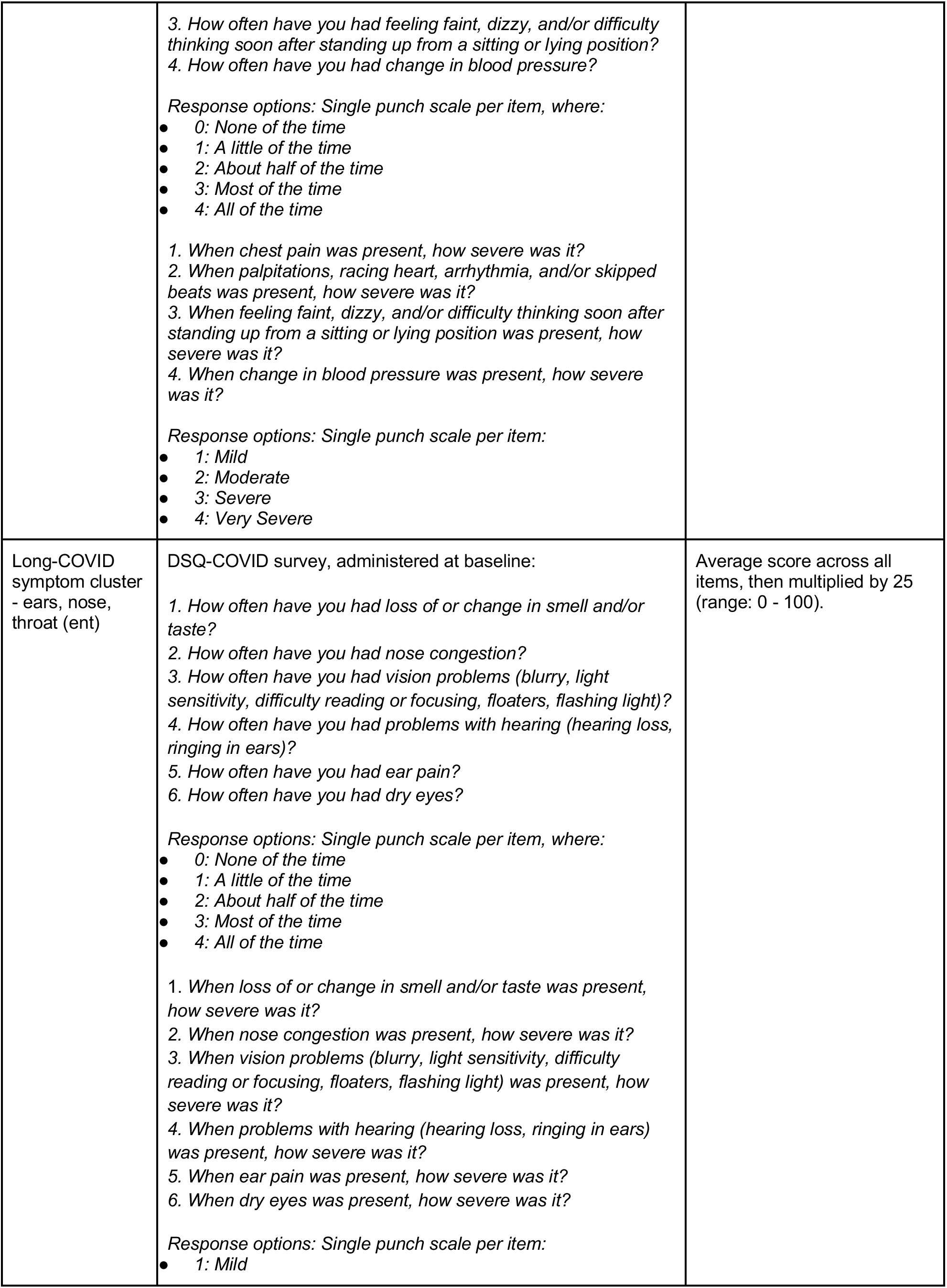
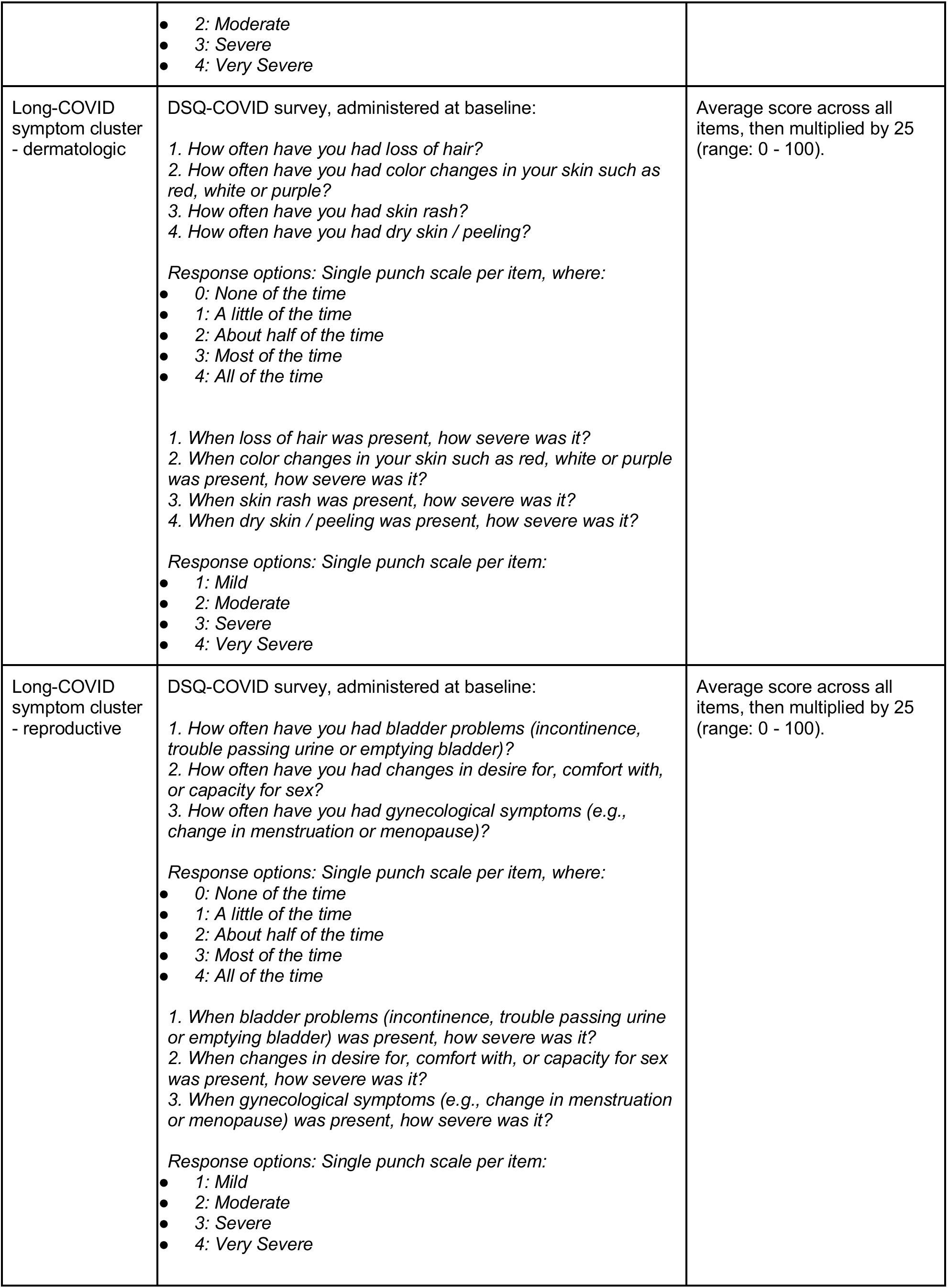
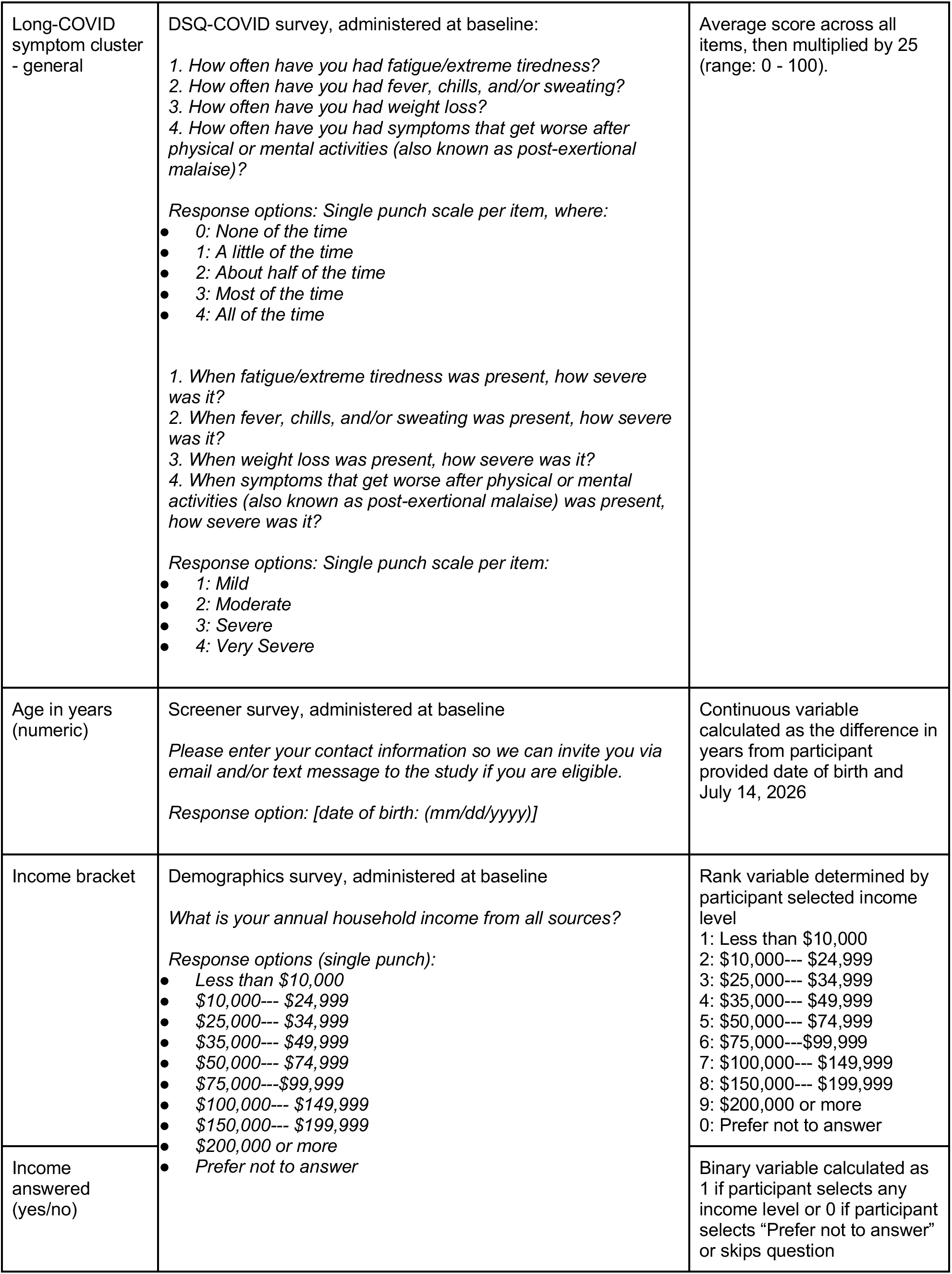
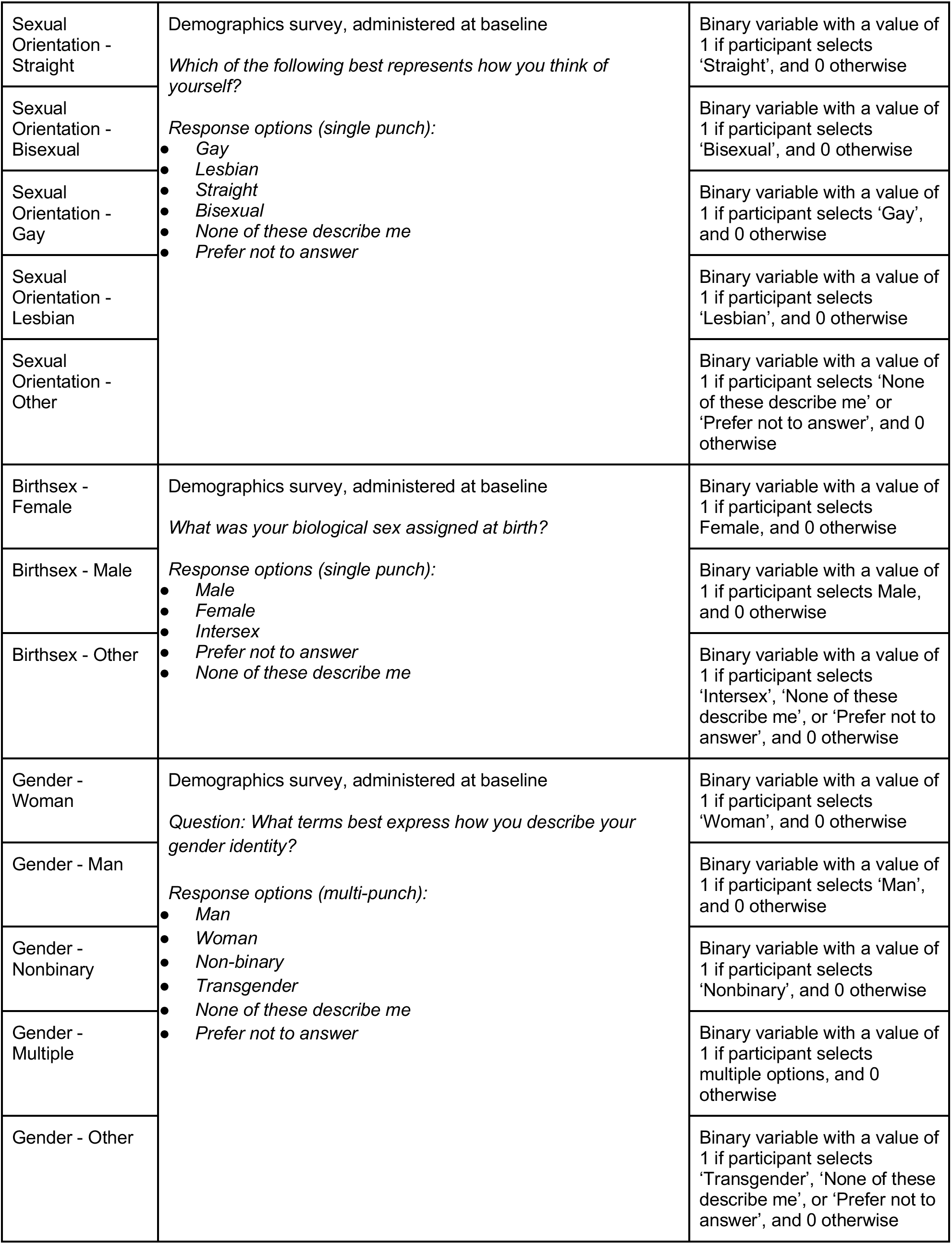
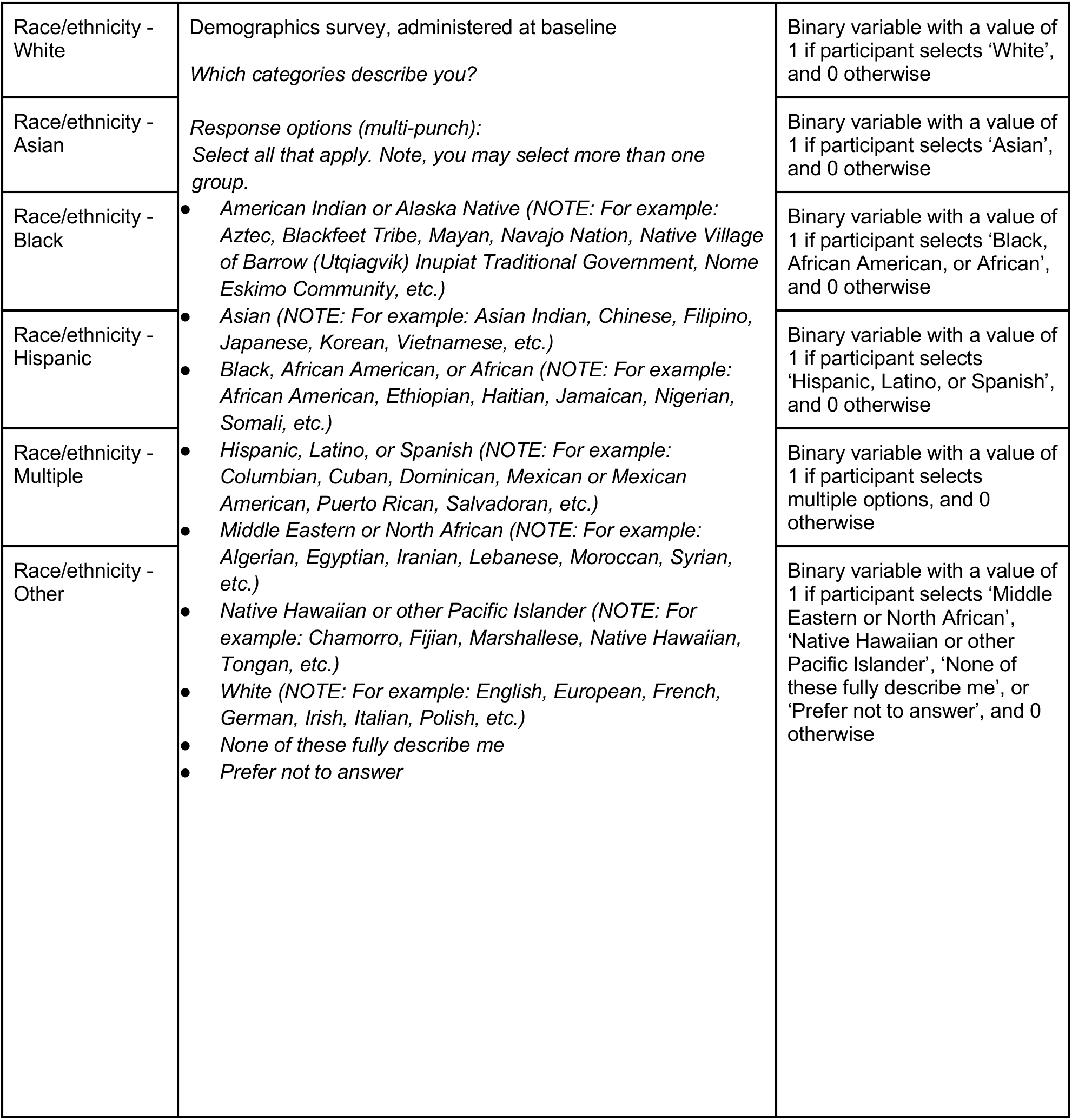
Data sources and operationalization of each control variable.

### 3. Participant Centric Design

**Figure S1.**
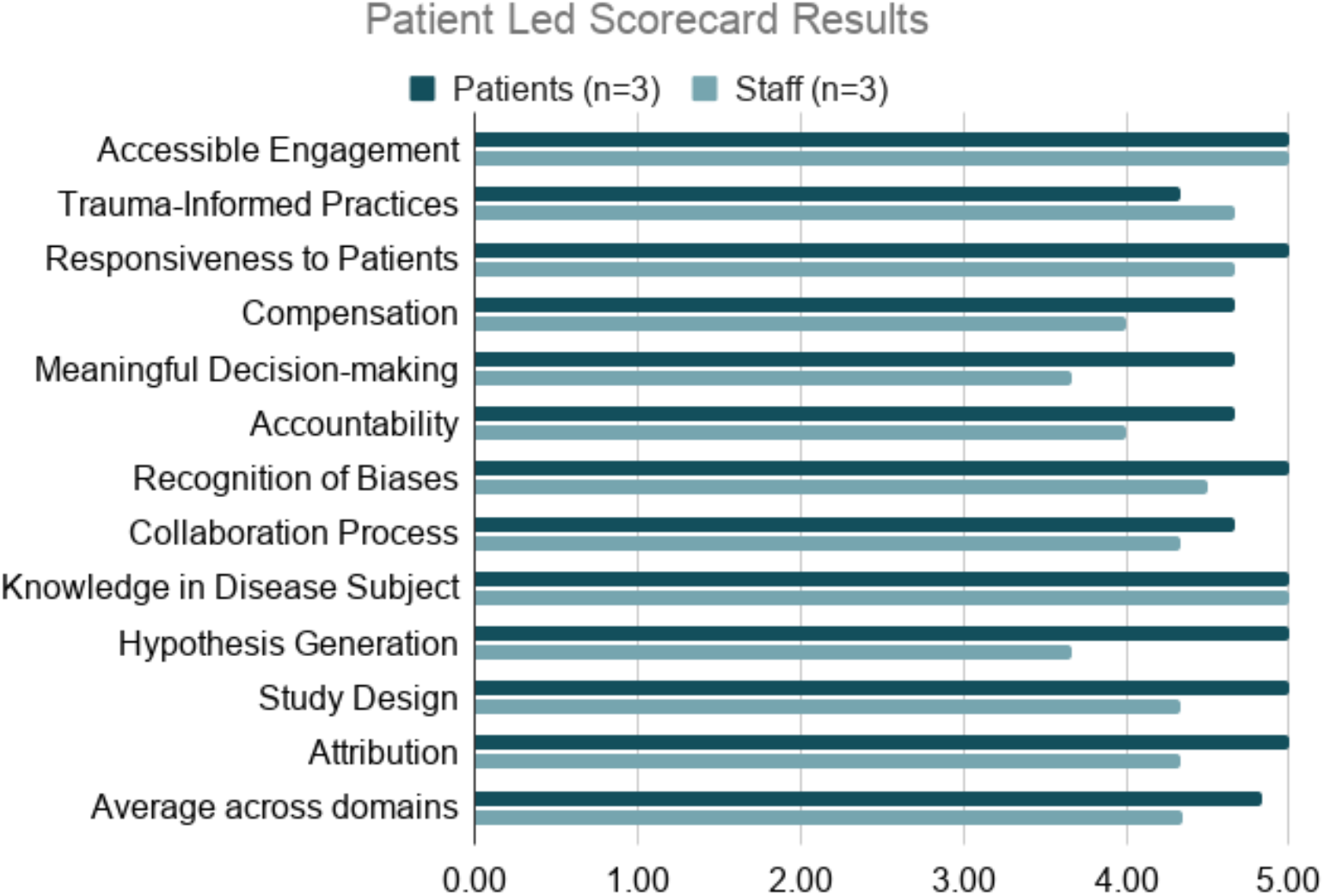
Patient Led Research Scorecard results.

### 4. Participant Communications

#### Participant communications processes and feedback

By default, participants are reminded (by email and/or SMS depending on their preferences) to complete study tasks up to three times. Some participants found the communication volume excessive. Following receipt of an adverse event, the study team only contacted participants if they had follow-up questions; some participants wanted personalized follow-up when their adverse event was reviewed and others did not. Through qualitative feedback, some participants requested more responses on weekends and after hours, at which times the team only responded to urgent matters. A similar pattern emerged with injection education; some participants were comfortable receiving the information in a webinar, some preferred individual calls, and some reported not needing any instruction from study staff. Finally, some participants indicated that the study team’s response times were delayed relative to their expectations. Going forward, we will offer more options to customize communication frequency and more clearly communicate expected communication timeframes.

#### Participant communications guidelines

**Table S4.** Communication guidelines.

|  |
| --- |
| Preferred language & tone |

| Category | Example phrases |
| --- | --- |
| Empathy & Respect | "We hope you are not experiencing any ongoing symptoms of COVID-19 but if you are, you are not alone." |
| Clear and concise (7th grade level) | "This is a research study and participation is completely up to you."<br>"We are studying if tirzepatide can help individuals with Long COVID feel better." |
| Accessibility | "Participation can be completed entirely from home, no in-person visits needed."<br>"MyDataHelps™ smartphone app by CareEvolution for remote participation, electronic health record sharing, connecting devices, completing surveys and more!"<br>"Our study team is available to support you in the way that works best for you, whether that's by phone, email or video call." |
| Hopeful, not misleading | "In the long-term, this clinical trial could help researchers find therapies that work for everyone experiencing Long COVID."<br>"You may even learn ways to manage your own health." |
| Transparency | "Taking part in LoCITT-T is up to you. If you join, you can leave the study at any time." |

Table S4. Communication guidelines
| Category | Reason | Example phrases |
| --- | --- | --- |
| Dismissive | Minimizes patient experience. Downplays the fatigue, cognitive difficulty and barriers people with Long COVID face, even from home. | "We want to understand why people *think* they still have symptoms of Long COVID."<br>"You don't have to leave home, so it's easy!" |
| Overpromising or misleading | Implies guaranteed outcomes | "This clinical trial will cure Long COVID."<br>"Join now for the best treatment." |
| Jargon or complexity | Confusing or inaccessible | "Randomized, placebo-controlled" (without further explanation) |
| Stigmatizing or invalidating | May trigger or alienate participants | “Unexplained symptoms”<br>“Alleged Long COVID” |
| Overly clinical or detailed | Lacks empathy | “Data will be extracted for research purposes.”<br>“Participants will be evaluated for compliance.” |

### 5. Comparison of LoCITT-T enrollment timelines against completed and ongoing Long COVID clinical trials

**Table S5.** Comparison of LoCITT-T against completed and ongoing Long COVID clinical trials.

| Trial ID | Enrollment status | Country | Target N | Enrollment period | Monthly accrual rate |
| --- | --- | --- | --- | --- | --- |
| PANORAMIC Norway [NCT05852873] | Closed | Norway | 2000 | 2023-05-12 – 2024-09-09 | 125.26 |
| RECLAIM [NCT07280572] | Open | Netherlands | 1250 | 2025-02-17 – 2027-09-08 | 40.78 |
| LOCITT-T * [NCT07128082] | Closed | United States | 1000 | 2025-10-24 – 2026-01-05 | 416.95 |
| RECOVER-SLEEP [NCT06404086] | Closed | United States | 830 | 2024-07-31 – 2024-12-31 | 165.12 |
| Synthetic Vitamin B1 [NCT05642923] | Closed | Egypt | 528 | 2023-01-08 – 2024-03-02 | 38.36 |
| NCT06404112 | Closed | United States | 470 | 2024-07-31 – 2025-12-25 | 27.94 |
| NCT05513560 | Closed | Canada | 460 | 2023-05-31 – 2025-01-29 | 22.99 |
| NCT05220280 | Closed | Finland | 400 | 2022-02-06 – 2022-12-02 | 40.72 |
| NCT06128967 | Closed | Brazil | 399 | 2023-10-18 – 2025-05-02 | 21.61 |
| NCT06383819 | Closed | Russian Federation | 392 | 2022-04-08 – 2023-10-21 | 21.27 |
| NCT06305780 | Closed | United States | 381 | 2024-03-11 – 2024-10-08 | 54.96 |
| NCT06404099 | Closed | United States | 361 | 2024-08-12 – 2026-01-06 | 21.46 |
| NCT06928272 | Closed | United States, Brazil, Canada, Italy, Uganda, Zambia | 348 | 2025-09-10 – 2026-06-01 | 40.12 |
| NCT05638633 | Closed | Germany | 321 | 2022-11-11 –<br>2025-01-02 | 12.48 |
| NCT06721949 | Open | Canada | 300 | 2025-11-18 –<br>2027-11-02 | 12.79 |
| NCT05619653 | Closed | Austria, Germany | 279 | 2022-12-12 –<br>2025-04-18 | 9.90 |
| NCT05481177 | Closed | United States | 250 | 2023-06-14 –<br>2024-06-02 | 21.50 |
| NCT05823896 | Closed | Sweden | 219 | 2023-05-01 –<br>2024-11-12 | 11.88 |
| NCT06847191 | Closed | United States | 203 | 2025-04-29 –<br>2026-05-09 | 16.48 |
| NCT05497089 | Closed | Italy, Spain,<br>Switzerland | 203 | 2022-08-29 –<br>2023-11-24 | 13.67 |
| NCT04978259 | Closed | Finland | 202 | 2021-07-24 –<br>2022-02-28 | 28.07 |
| NCT06305793 | Closed | United States | 200 | 2024-03-11 –<br>2025-06-22 | 13.01 |
| NCT06590324 | Closed | Jordan, Saudi<br>Arabia, United<br>Arab Emirates | 200 | 2025-04-15 –<br>2025-10-01 | 36.02 |
| NCT05874037 | Closed | United States | 191 | 2023-05-15 –<br>2024-11-09 | 10.69 |
| NCT04904536 | Closed | Australia | 190 | 2022-03-10 –<br>2024-06-30 | 6.86 |
| NCT05926505 | Closed | Germany, Greece,<br>Italy, Spain | 182 | 2023-09-06 –<br>2026-03-02 | 6.10 |
| NCT06305806 | Closed | United States | 181 | 2024-03-11 –<br>2025-07-14 | 11.24 |
| NCT06643299 | Closed | United States | 180 | 2025-05-13 –<br>2025-08-30 | 50.26 |
| NCT06366724 | Closed | United States | 160 | 2024-09-10 –<br>2026-05-19 | 7.91 |
| NCT05430152 | Closed | Canada | 160 | 2024-01-15 –<br>2025-10-26 | 7.49 |
| NCT05890534 | Closed | Switzerland | 153 | 2023-06-07 –<br>2024-08-13 | 10.76 |
| NCT05684952 | Closed | China | 152 | 2023-05-30 –<br>2023-12-18 | 22.90 |
| NCT05795816 | Closed | Australia | 150 | 2023-10-25 –<br>2024-11-07 | 12.05 |
| NCT05212610 | Closed | United States | 137 | 2022-03-21 –<br>2025-02-28 | 3.88 |
| NCT05967052 | Open | Poland | 132 | 2023-10-24 –<br>2027-08-31 | 2.86 |
| NCT05911009 | Closed | Austria, Finland,<br>Germany, Spain,<br>Switzerland | 119 | 2023-06-16 –<br>2024-05-27 | 10.47 |
| NCT06792214 | Closed | Canada | 118 | 2025-01-03 –<br>2025-01-01 | 9.84 |
| NCT04944121 | Closed | United States | 112 | 2021-06-25 –<br>2023-08-21 | 4.33 |
| NCT05747534 | Closed | United States | 107 | 2023-05-31 –<br>2026-04-10 | 3.12 |
| NCT05697640 | Closed | Germany | 104 | 2023-06-22 –<br>2026-01-21 | 3.35 |
| NCT05524532 | Closed | United States,<br>Puerto Rico | 101 | 2023-07-20 –<br>2025-05-06 | 4.69 |
| NCT07021794 | Open | United States | 100 | 2025-06-16 –<br>2026-07-16 | 7.71 |
| NCT06441955 | Open | United States | 100 | 2024-03-01 –<br>2027-09-30 | 2.33 |
| NCT06980636 | Open | China | 100 | 2025-12-01 –<br>2026-08-07 | 12.22 |
| NCT05668091 | Closed | United States | 100 | 2023-04-14 –<br>2024-03-12 | 9.14 |
| NCT07093580 | Closed | United States | 98 | 2025-07-29 –<br>2026-03-04 | 13.68 |
| NCT06766825 | Closed | Spain | 90 | 2025-02-07 –<br>2026-03-03 | 7.04 |
| NCT06511063 | Closed | United States | 90 | 2024-10-01 –<br>2026-05-05 | 4.71 |
| NCT07627815 | Open | Canada | 82 | 2026-05-20 –<br>2026-07-15 | 44.57 |
| NCT06960928 | Open | United States | 80 | 2025-04-18 –<br>2026-07-16 | 5.36 |
| NCT07298005 | Open | Netherlands | 80 | 2026-04-01 –<br>2026-11-02 | 11.33 |
| NCT04842448 | Closed | Sweden | 80 | 2021-09-15 –<br>2023-06-28 | 3.74 |
| NCT06492798 | Closed | China | 76 | 2023-09-01 –<br>2025-06-03 | 3.61 |
| NCT05911906 | Closed | United Kingdom | 73 | 2024-10-08 – 2025-07-24 | 7.69 |
| NCT06055244 | Closed | United States | 64 | 2023-12-07 – 2025-08-11 | 3.18 |
| NCT05472090 | Closed | United States | 63 | 2022-08-18 – 2023-04-05 | 8.34 |
| NCT06204432 | Closed | United States | 60 | 2023-12-01 – 2024-12-01 | 4.99 |
| NCT06316843 | Closed | United States | 59 | 2023-10-15 – 2024-08-08 | 6.03 |
| NCT06159283 | Closed | Korea, Republic of | 58 | 2024-03-18 – 2025-06-16 | 3.88 |
| NCT06419712 | Closed | Mexico | 54 | 2022-11-25 – 2023-02-18 | 19.34 |
| NCT05618587 | Closed | United States | 52 | 2022-11-28 – 2023-06-30 | 7.40 |
| NCT07189936 | Open | United States | 50 | 2025-12-18 – 2028-06-02 | 1.70 |
| NCT04482595 | Closed | United States | 50 | 2020-11-11 – 2024-05-08 | 1.19 |
| Note: The enrollment period is determined using two dates available from clinicaltrials.gov. First, we take the “Study start” date (Actual or Estimated) to mark the start of the enrollment period. Second, we infer the enrollment end date using the date reported under “primary completion (estimated)” and subtract the observation period listed under the primary outcome. If multiple timeframes are listed, the longest timeframe is used. We then calculate the monthly enrollment date by dividing the “Enrollment” (Actual or Estimated) by the number of days between the end and start for the enrollment period, multiply by 365.25 and divide by 12.<br>* For LOCITT-T (NCT07128082), we manually overrode the enrollment dates to 10/24/2025 - 01/05/2026 to give a more conservative estimate. In particular, the declared end date on clinicaltrials.gov reflected the closing of the eligibility survey, not when we enrolled the first participant. Further, the declared start date excluded testing of family and friends. |  |  |  |  |  |

#### Methods for Comparison of LoCITT-T enrollment timelines against completed and ongoing Long COVID clinical trials

##### Data extraction process

We use publicly available information from clinicialtrial.gov to build a dataset of Long COVID trials. First, we extract a list of trials using the clinicialtrial.gov API using a query for studies where the ‘condition’ = ‘Long Covid OR “Post-COVID Syndrome” OR “Post-COVID-19 Syndrome” OR “Post-Acute Sequelae of SARS-CoV-2 Infection’; AND ‘study type’ = ‘interventional’. This yields a list of 358 studies.

##### Exclusion criteria

We then apply several exclusions. First, we exclude trials where the intervention is not pharmaceutical, operationalized as including mention of the word ‘drug’ under the ‘intervention/treatment’ field on the clinicialtrial.gov webpage for the trial. This information was extracted using ChatGPT 5.5 (OpenAI, June 12, 2026). This exclusion reduces the list to 89 trials. Following a manual inspection, we further excluded 11 trials that were not specific to Long COVID symptoms, consisted of a diagnostic investigation, or targeted the incidence or effects of COVID-19 infection, rather than Long COVID. This reduces the list to 78 trials. Finally, we apply an exclusion criterion requiring the trial to have an enrollment (actual or estimated) of at least 50 participants, which we believe makes the results on monthly accrual rate more interpretable. This further excludes 15 trials, leaving a final list of 63.

##### Calculation of the enrollment period and monthly enrollment rate

The enrollment period is determined using two dates available from clinicialtrials.gov. First, we take the “Study start” date (Actual or Estimated) to mark the start of the enrollment period. Second, we infer the enrollment end date using the date reported under “primary completion (estimated)” and subtract the observation period listed under the primary outcome. If multiple timeframes are listed, the longest timeframe is used. We then calculate the monthly enrollment date by dividing the “Enrollment” (Actual or Estimated) by the number of days between the end and start for the enrollment period, multiplied by 365.25 and divided by 12.

For LOCITT-T, we manually overrode the enrollment dates to 10/24/2025 - 01/05/2026 to give a more conservative estimate. In particular, the declared end date on clinicialtrials.gov reflected the closing of the eligibility survey, not when we enrolled the first participant. Further, the declared start date excluded testing of family and friends.

##### AI prompt

Each row in this spreadsheet contains information about a study. Please do the following: * Using the identifier in column “nctid”, find the study on clinicialtrial.gov using the URL “https://clinicaltrials.gov/study/[nctid]”. For example you can find study with ID “NCT05852873” at https://clinicaltrials.gov/study/NCT05852873 * From the website, grab the following: Sponsor, Information provided by, Study Start (Actual or Estimated, whichever is relevant), Primary Completion (Actual or Estimated), Enrollment (Actual or Estimated), Study Type, Phase, Arms and Interventions, Primary Outcome Measures, Measure Description for each Primary Outcome Measure, Time Frame for Primary Outcome Measure * Populate this information in a spreadsheet. * Add a column called Clinical Trial and another column for Rationale. Please populate Clinical Trial with a value of ‘yes’ if you think this trial is a clinical trial and ‘no’ otherwise. Use the Rationale column to provide a rationale for why you think so. * Add a column called “Completed Enrollment” and put a date there that is calculated as follows: Take the date under Primary Completion (Actual) or Primary Completion (Estimated) and subtract the timeframe listed under the Primary Outcome Measure. For example, if the timeframe listed is 12 weeks, then subtract 12 weeks from Primary Completion date and report that under Completed Enrollment. If multiple timeframes are reported, take the longest one. * Then, add a column Completed Enrollment Actual/Estimated. Populate the cell with “Actual” if the date under Completed Enrollment is in the past, and Estimated if it is today or in the future. * Add a column called Enrollment Days that is the number of days between Completed Enrollment and Study Start * Add a column called Monthly Enrollment Rate that is the Enrollment number divided by Enrollment Days * 365.25 divided by 12.

Please amend the spreadsheet by adding two additional columns: * Pharmaceutical Intervention to capture whether the intervention is a drug or medication. Lifestyle changes, exercise, or diet, are not considered pharmaceutical interventions. You can add a column with rationale. Note that if one of the intervention arms includes a pharmaceutical intervention, it can be included. * Add a Country column to represent the location of the trial. If multiple countries are listed, please use additional columns.

### 6. Comparison of LoCITT-T total and per-participant costs against other clinical trials

We sought to compare the total and per-participant costs of LoCITT-T to other drug clinical trials. First, we established the list of trials by searching NIH RePORTER with the term “clinical trial,” resulting in a list of over 15,000 trials. Next we used AI (Claude) to search ClinicalTrials.gov for information on enrollment volume for all trials that were not phase 1 or first in human, with a budget of at least $2.5M; this cost restriction was a practicality because the volume of trials at lower costs is much higher and the participant number is expected to be lower. We also prompted the AI to search for any additional trial cost data. When data was available on at least half of the project years and enrollment volume, the trial was included in the cost per participant calculations. The AI-populated list consisted of a total of 55 trials.

Next, the sources for cost and enrollment volume for all included trials were manually reviewed by two of the manuscript authors and 43 (78.2%) were updated for accuracy; in most cases not all years of funding had been included to generate the total costs. To manually verify cost information, we searched the project number in NIH RePORTER, went to the “History” section of the NIH RePORTER page and added the listed costs. In most cases the Clinical Studies section of the project description contained a link to the ClinicalTrials.gov page; in some cases where there was not a direct link, a search was performed for a trial with a matching intervention, Principal Investigator, and trial description. The manual review led to the exclusion of four trials. For two, matching trial data could not be found; for other two trials, NIH RePORTER only listed funding information for an extension period. This left 51 trials that were included in the final analysis.

RECOVER trials were reported on in aggregate based on the publicly announced amount of the first phase of clinical trials, dividing it by the total number of participants enrolled in those trials based on ClinicalTrials.gov. Note that for ongoing trials, the full costs may be an undercount since their full costs are not yet available. The file containing this data is included as supplemental data.

